# Highly comparative time series analysis of oxygen saturation and heart rate to predict respiratory outcomes in extremely preterm infants

**DOI:** 10.1101/2024.01.24.24301724

**Authors:** Jiaxing Qiu, Juliann M. Di Fiore, Narayanan Krishnamurthi, Premananda Indic, John L. Carroll, Nelson Claure, James S. Kemp, Phyllis A. Dennery, Namasivayam Ambalavanan, Debra E. Weese-Mayer, Anna Maria Hibbs, Richard J. Martin, Eduardo Bancalari, Aaron Hamvas, J. Randall Moorman, Douglas E. Lake

**Affiliations:** Department of Medicine, Division of Cardiology, University of Virginia School of Medicine, Charlottesville, VA; Department of Pediatrics, Case Western Reserve University School of Medicine, University Hospitals Rainbow Babies and Children’s Hospital, Cleveland, OH; Department of Pediatrics, Division of Autonomic Medicine, Northwestern University Feinberg School of Medicine, Chicago, IL; Department of Pediatrics, Division of Neonatology, University of Miami Miller School of Medicine, Miami, FL; Department of Pediatrics, Division of Neonatology, University of Alabama at Birmingham, Birmingham, AL; Department of Pediatrics, Division of Newborn Medicine, Washington University School of Medicine, St. Louis, MO; Department of Pediatrics, Brown University School of Medicine, Department of Pediatrics, Providence, RI; Department of Electrical Engineering, University of Texas at Tyler, Tyler, TX; Ann and Robert H. Lurie Children’s Hospital and Northwestern University Department of Pediatrics, Chicago, IL; Department of Pediatrics, Division of Pediatric Pulmonology, Washington University School of Medicine, St. Louis, MO; Department of Pediatrics, University of Arkansas for Medical Sciences and Arkansas Children’s Hospital, Little Rock, AK

**Keywords:** highly comparative time series analysis, preterm infants, intermittent hypoxemia, predictive models

## Abstract

**Objective:** Highly comparative time series analysis (HCTSA) is a novel approach involving massive feature extraction using publicly available code from many disciplines. The Prematurity-Related Ventilatory Control (Pre-Vent) observational multicenter prospective study collected bedside monitor data from *>* 700 extremely preterm infants to identify physiologic features that predict respiratory outcomes. We calculated a subset of 33 HCTSA features on *>* 7*M* 10-minute windows of oxygen saturation (SPO2) and heart rate (HR) from the Pre-Vent cohort to quantify predictive performance. This subset included representatives previously identified using unsupervised clustering on *>* 3500 HCTSA algorithms. Performance of each feature was measured by individual area under the receiver operating curve (AUC) at various days of life and binary respiratory outcomes. These were compared to optimal PreVent physiologic predictor IH90 DPE, the duration per event of intermittent hypoxemia events with threshold of 90%.

**Main Results:** The top HCTSA features were from a cluster of algorithms associated with the autocorrelation of SPO2 time series and identified low frequency patterns of desaturation as high risk. These features had comparable performance to and were highly correlated with IH90 DPE but perhaps measure the physiologic status of an infant in a more robust way that warrants further investigation. The top HR HCTSA features were symbolic transformation measures that had previously been identified as strong predictors of neonatal mortality. HR metrics were only important predictors at early days of life which was likely due to the larger proportion of infants whose outcome was death by any cause. A simple HCTSA model using 3 top features outperformed IH90 DPE at day of life 7 (.778 versus .729) but was essentially equivalent at day of life 28 (.849 versus .850). These results validated the utility of a representative HCTSA approach but also provides additional evidence supporting IH90 DPE as an optimal predictor of respiratory outcomes.

## 1. Introduction

### 1.1. Highly comparative time series analysis (HCTSA)

Analysis of heart rate (HR) and oygen saturation (SPO2 or more precisely *SpO*_2_) vital sign time series during the stay of infants in the neonatal intensive care unit (NICU) has been shown to be useful in predicting unfavorable outcomes. This is part of larger efforts to improve care of infants using big data analytics (Vesoulis et al. 2023, Cole 2021). Notably, models for predicting sepsis have not only been developed (Griffin et al. 2003, Fairchild et al. 2017, Kausch et al. 2023) but implemented in the NICU and shown to improve outcomes in a large clinical trial (Moorman et al. 2011, Fairchild et al. 2013). Models have also been developed for other adverse outcomes during NICU stay including death (Sullivan et al. 2016, 2018, Niestroy et al. 2022), bronchopulmonary dysplasia (BPD) (Raffay et al. 2019, Gentle et al. 2023, Ramanand et al. 2023), and retinopathy of prematurity (Di Fiore et al. 2010, 2012). Vital sign data during an infant’s stay has also been shown to predict long-term outcomes of cognitive impairment (Poets et al. 2015), cerebral palsy (Letzkus et al. 2022), and autism (Blackard et al. 2021).

Highly comparative time-series analysis (HCTSA) developed by Fulcher et al. (2013) is a novel method that naturally applies to HR and SPO2 data. The core concept involves using numerous time-series algorithms with a wide-ranging set of parameter values to extract a massive number of features to associate with some target outcome. They examined over 35,000 real-world and model-generated time series with more than 7,000 time-series analysis algorithms developed from a wide variety of disciplines. The MATLAB code to implement these HCTSA algorithms is publicly available at https://github.com/benfulcher/hctsa. The goal of using HCTSA is not necessarily to develop an optimal predictive model restricted to these algorithms. Here we use HCTSA as a tool to identify new types of algorithms for future more traditional development with sufficient understanding to be useful for clinicians.

Recently, Niestroy et al. (2022) applied *>* 3500 HCTSA algorithms to *>* 17M 10-minute windows of heart rate (HR) and oxygen saturation (SPO2) vital sign data collected from bedside monitors (displayed every two seconds) from 6000 infants at the University of Virginia NICU from 2009 to 2019. The data (including all HR and SPO2 time series) is publicly available at Niestroy et al. (2021). In an effort to reduce the high dimensionality of the data, a random subset of *>* 120*K* daily results was then used for unsupervised k-medoids clustering based on distance metric of mutual information. With *k* = 20 clusters, 81% of the variance of full data was explained and identified 20 central algorithms or medoids. A medoid is defined to be a point in the cluster from which the sum of distances to other data points in the cluster is minimal.

Utilizing these clusters, they found that HCTSA algorithms can discover novel patterns associated with neonatal mortality in the next 7 days. Notably, models based solely on the cluster centers performed comparably to those considering the full feature set. This lead to a hypothesis that identifying algorithms by unsupervised clustering could capture most of the predictive information in NICU vital sign data and that motivates this work.

Based on this hypothesis, a subset of HCTSA algorithms (including at least one from each of the 20 clusters) was implemented using the publicly available code as part of the Batch Algorithm Processor (BAP) software package developed for comprehensive analysis of bedside waveform and vital sign time series data. In addition, the maximum and minimum cross-correlation of HR and SPO2 at lags up to 30 seconds were calculated which have been shown to be associated with sepsis, apnea and periodic breathing (Fairchild & Lake 2018) giving a total of 33 vital sign features calculated. The BAP software including extensive documentation is available at https://github.com/UVA-CAMA/BatchAlgorithmProcessor.

### 1.2. PreVent Study

The Prematurity-Related Ventilatory Control (Pre-Vent) study was of a cohort of *>* 700 extremely premature infants (gestational age less than 29 weeks) across 5 NICU sites with the hypothesis that physiologic features of ventilatory control extracted from bedside monitoring data can predict unfavorable respiratory outcomes at 40 weeks post-menstrual age (PMA) (Dennery et al. 2019). The BAP was developed for Pre-Vent and used to extract a large number of physiologic (including HCTSA) features. The software was run at each of the sites remotely in a separate but uniform way while keeping raw data stored locally. A result of this processing was 33 features calculated on 7.8*M* 10-minute windows of HR and SPO2 data.

In the primary analysis of the Pre-Vent study, optimal logistic regression models were developed for predicting respiratory outcomes at varying days of life during the NICU stay and identified important risk factors for both physiologic and clinical models (Ambalavanan et al. 2023). Physiologic models included metrics calculated from bedside waveforms (ECG and chest impedance) and vital signs (HR and SPO2). Notably, the duration per event (DPE) of intermittent hypoxemia events with SPO2*<* 90% (IH90) was most significant individual physiologic factor for predicting the primary unfavorable respiratory outcome. At day of life 28, performance of IH90 DPE as an individual predictor performed similarly to optimal models with demographic, respiratory support and other clinical features. However, IH and physiologic models in general did not perform as well earlier in the NICU stay (day 7) where identification of higher-risk infants would have more impact.

### 1.3. Intermittent Hypoxemia

Intermittent hypoxemia (IH) in the NICU has been widely studied (Ramirez et al. 2023, Dormishian, Schott, Aguilar, Jimenez, Bancalari, Tolosa & Claure 2023, Dormishian, Schott, Aguilar, Bancalari & Claure 2023). Definitions of IH events require fixed parameters including SPO2 threshold (e.g. 80% or 90%), min/max duration, and possibly joining rules for nearby events. These definitions can be somewhat arbitrary and susceptible to varying hospital protocols (e.g., target SPO2 ranges) or clinical practice.

IH events can also be sensitive to the averaging time of the pulse oximeter which can vary across vendor and NICU. Published physiological models relying on threshold-based events may not prove to be robust in universal and evolving applications. Using advanced time-series metrics to predict neonatal respiratory outcomes without relying on clinical definitions have not yet been fully studied.

## 2. Methods

### 2.1. Study Population

The study population consisted of the 717 extremely premature infants prospectively enrolled in Pre-Vent study and with respiratory outcome determined (Ambalavanan et al. 2023). Analysis was restricted to days where sufficient HR and SPO2 monitor data was available (at least 12 hours of each) which represented over 80 percent of the study period.

The Pre-Vent study predefined 5 mutually exclusive respiratory outcome categories which in order of decreased severity are:

i. **Death:** prior to 40 weeks PMA
ii. **Severe:** invasive mechanical ventilation (IMV) at 40 weeks PMA
iii. **Moderate:** need for positive pressure at 40 weeks PMA
iv. **Mild:** respiratory medications, oxygen or other respiratory support either inpatient at 40 weeks PMA or at discharge prior to 40 weeks PMA
v. **Favorable:** none of the above

The primary outcome of the Pre-Vent study was an unfavorable outcome consisting of any of the first 4 outcomes (i.e., not favorable). Three additional binary outcomes analyzed here in order of increase severity are moderate or severe or death, severe or death and death. For each outcome, the entire cohort was used to compare having vs. not having outcome.

Predictive performance of the time series features were evaluated at day of life 7, 14, and 28 which follows the approach of the PreVent analysis (Ambalavanan et al. 2023). Table 1 shows a breakdown of the infants by binary outcomes at these time points. A particular focus was predicting an unfavorable outcome at day 7, which consisted of 584 infants and an event rate of 265/584=45.4%. An important aspect of the analysis to consider is the evolving distribution of the outcomes by day of life. Figure 1 shows how the mortality rate of surviving infants is significantly reduced by week of life. Since death outcomes include those of all causes and not necessarily related to respiratory failure, analyzing outcomes later in the NICU stay are therefore more directly associated with control of breathing. Also note in Figure 1 that there is minimal difference in mortality rate between those with monitor data and entire population suggesting there is no significant selection bias in the analysis.

**Figure 1:**
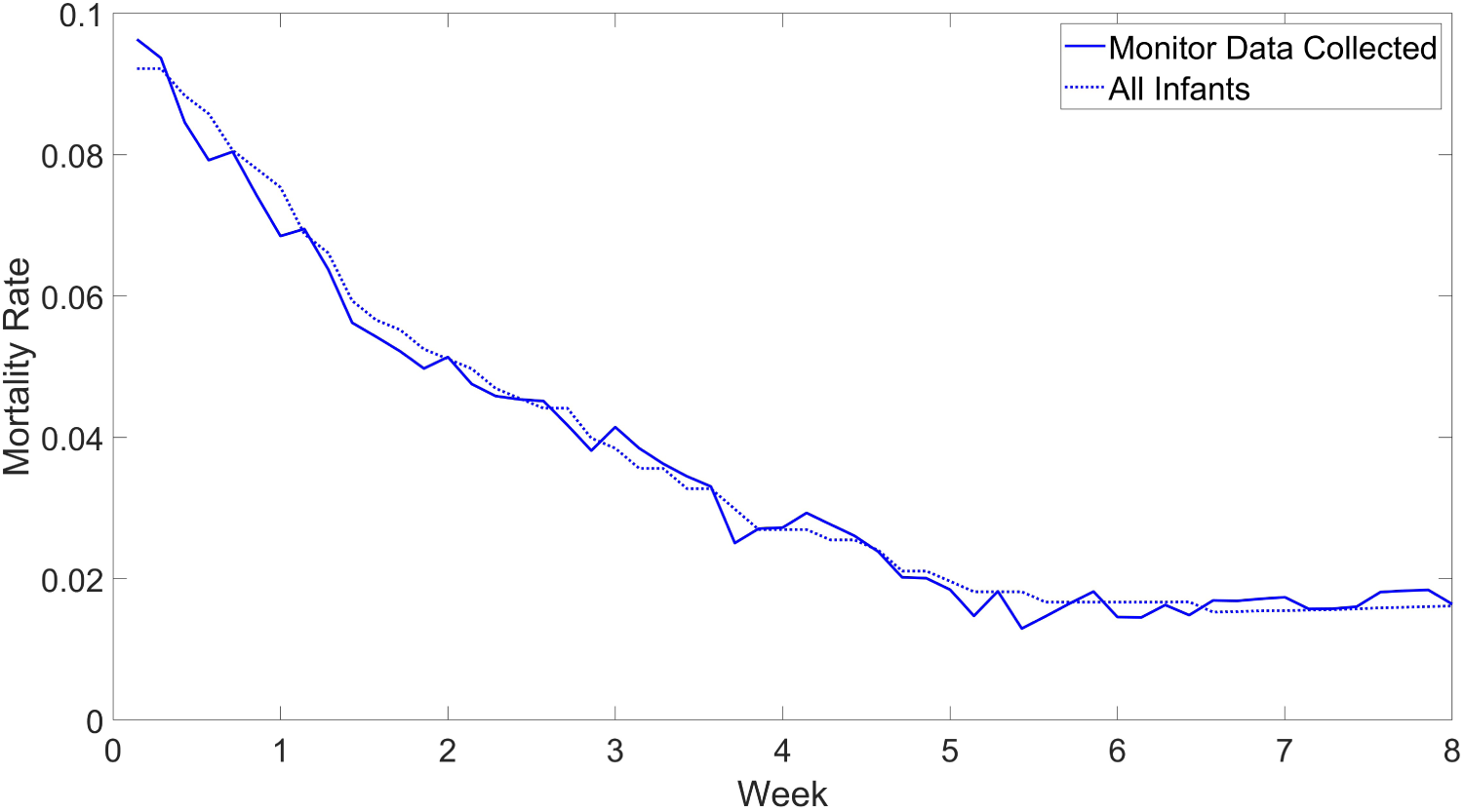
Mortality rate by week of life. For each time point, the solid line shows the mortality rate in patients with monitoring data collected and the dotted line shows the mortality rate in all infants in study.

**Table 1:**
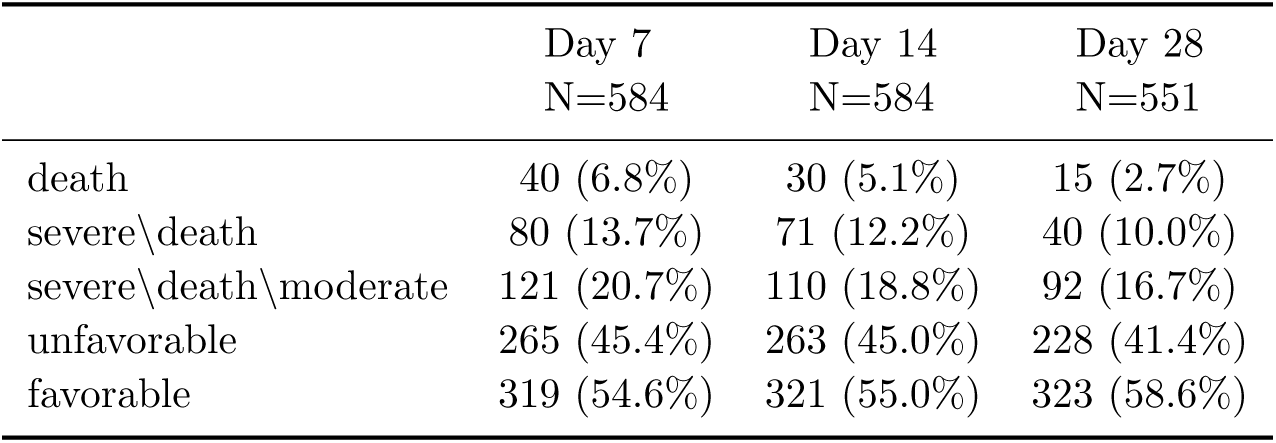
Sample size and distribution of binary respiratory outcome at day 7, 14, and 28 for infants with monitor data collected.

### 2.2. Time Series Features

The 33 new HR and SPO2 HCTSA features are described in Table 2 along with which of the 20 clusters they belonged to and a general description of the type of algorithms in the cluster. The number of the clusters are ordered in way so that nearby clusters tend to be more related. Table A1 provides more details on the exact parameters and the publicly available Matlab code used for each feature. For evaluation purposes, the features were summarized as the daily median of the every 10 minute results.

**Table 2:**
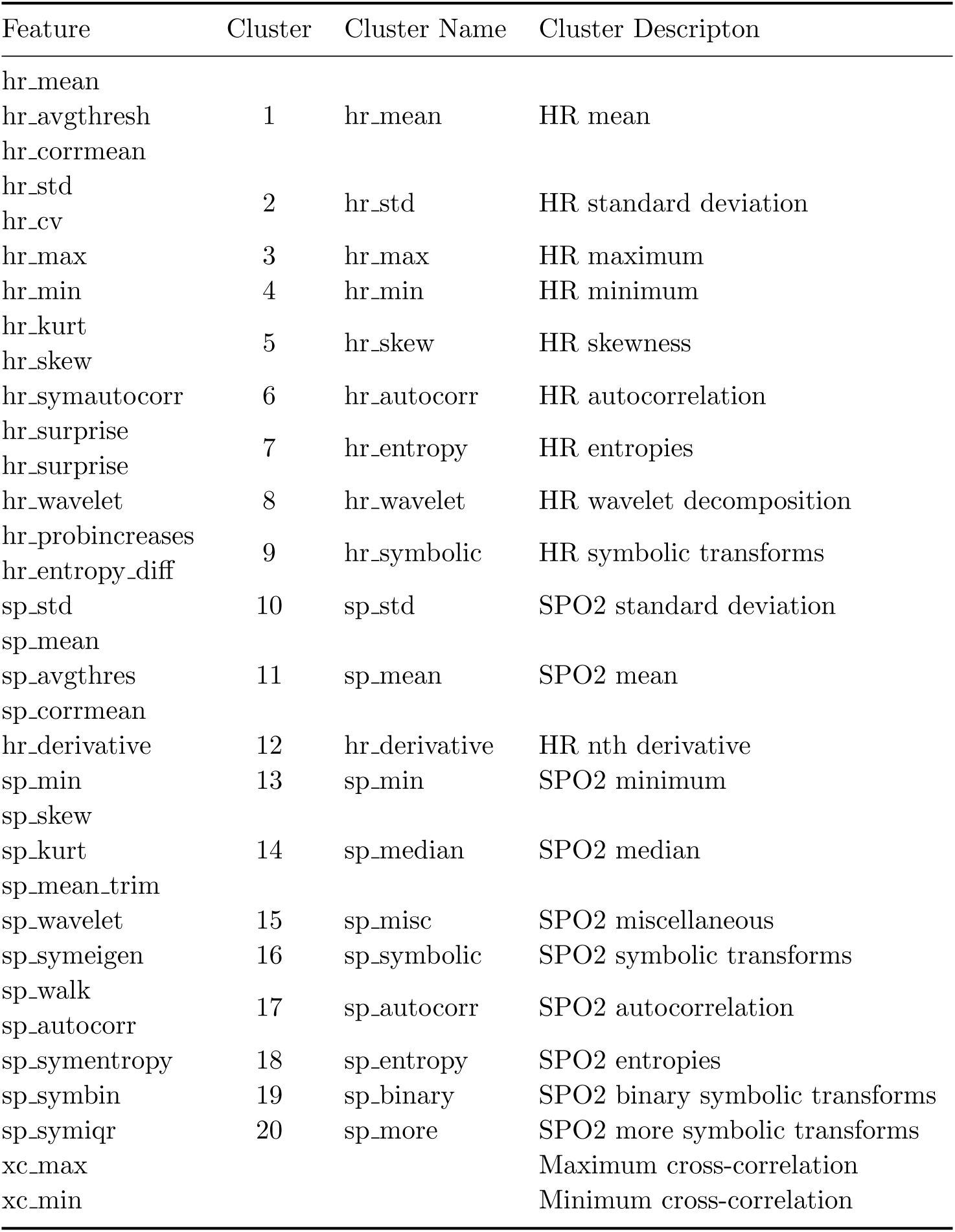
Time-series features implemented in Batch Algorithm Processor (BAP).

| Feature | Cluster | Cluster Name | Cluster Description |
| --- | --- | --- | --- |
| hr_mean | 1 | hr_mean | HR mean |
| hr_avgthresh |  |  |  |
| hr_corrmean |  |  |  |
| hr_std | 2 | hr_std | HR standard deviation |
| hr_cv |  |  |  |
| hr_max | 3 | hr_max | HR maximum |
| hr_min | 4 | hr_min | HR minimum |
| hr_kurt | 5 | hr_skew | HR skewness |
| hr_skew |  |  |  |
| hr_symautocorr | 6 | hr_autocorr | HR autocorrelation |
| hr_surprise | 7 | hr_entropy | HR entropies |
| hr_surprise |  |  |  |
| hr_wavelet | 8 | hr_wavelet | HR wavelet decomposition |
| hr_probincreases | 9 | hr_symbolic | HR symbolic transforms |
| hr_entropy_diff |  |  |  |
| sp_std | 10 | sp_std | SPO2 standard deviation |
| sp_mean | 11 | sp_mean | SPO2 mean |
| sp_avgthres |  |  |  |
| sp_corrmean |  |  |  |
| hr_derivative | 12 | hr_derivative | HR nth derivative |
| sp_min | 13 | sp_min | SPO2 minimum |
| sp_skew | 14 | sp_median | SPO2 median |
| sp_kurt |  |  |  |
| sp_mean_trim |  |  |  |
| sp_wavelet | 15 | sp_misc | SPO2 miscellaneous |
| sp_symeigen | 16 | sp_symbolic | SPO2 symbolic transforms |
| sp_walk | 17 | sp_autocorr | SPO2 autocorrelation |
| sp_autocorr |  |  |  |
| sp_symentropy | 18 | sp_entropy | SPO2 entropies |
| sp_symbin | 19 | sp_binary | SPO2 binary symbolic transforms |
| sp_symiqr | 20 | sp_more | SPO2 more symbolic transforms |
| xc_max |  |  | Maximum cross-correlation |
| xc_min |  |  | Minimum cross-correlation |

The daily HCTSA results were merged with the physiological features measured on HR and SPO2 as part of PreVent study. Using the BAP, the following physiologic events were extracted from bedside sign monitor data:

i. IH80: Desaturation events with SPO2 *<* 80% for 10-300 seconds
ii. IH90: Desaturation events with SPO2 *<* 90% for 10-300 seconds
iii. Brady80: Bradycardia events with HR *<* 80 beats per minute for 5 or more seconds

These IH duration limits come from Di Fiore et al. (2012) but do vary slightly across publications. For each of these 3 events, daily features were quantified as count of events per day (count), total daily duration in events (dur) measured in minutes per day, and average duration per event (dpe) measured in seconds per event. These 3 quantities are not independent and have relationship dpe=60*×*dur/count. These 9 clinically defined features gave a total of 42 candidate HR and SPO2 metrics for the analysis.

### 2.3. Feature Evaluation

We evaluated the performance of each feature based on the area under the receiver operating curve (AUC) at various days of life for all 4 binary outcomes. The AUC for predicting the primary unfavorable outcome at day 7 was used to rank the features. Signature of risk curves showing estimates of the probability of the unfavorable outcome at day 7 as a function of the daily median value of each variable were made using logistic regression model including a cubic spline transformation with 3 knots to account for possible non-linearity. The top performers were then analyzed further by looking at a range of examples stratified by percentile to better understand what the algorithm was measuring and, in particular, its correlation with IH events. The trajectories of the performance of these new metrics was also analyzed for the first 8 weeks of life.

For logistic regression models, AUC was measured using cross-validation where each fold consisted of infants clustered by pregnancy so that an infant’s sibling was not used to predict its outcome. Variable importance was quantified by the drop in cross-validated AUC when the variable was removed from the model.

### 2.4. Demographic and Clinical Risk Factors

Detailed demographic information about the Pre-Vent cohort can be found in Ambalavanan et al. (2023). The sex of the infants in the study were equally represented (51% male) and the average birth weight and gestational age were 871 grams and 26.4 weeks rspectively. There is also a comprehensive list of clinical risk factors for predicting respiratory outcomes at various time points. Based on this, the following were identified as the major risk factors for unfavorable respiratory outcomes and included in the presented results for comparison purposes.

i. BW: birthweight in grams
ii. GA: gestational age in weeks
iii. IMV: daily need for invasive mechanical ventilation
iv. FIO2 (or *FiO*_2_): value closest to noon (0.21 if on room air)

We acknowledge the importance of demographic and clinical risk factors for predicting respiratory outcomes, but the focus of this paper is on prediction using only vital signs HR and SPO2. We can justify this as still being relevant clinically for a couple of reasons. First, vital sign features are dynamic and indicate the evolving status of the infant during the NICU stay whereas demographic features are static and fixed at birth. Second, clinical features like those associated with respiratory support are often in response to changes in the vital signs of the infant and potentially not a direct informative predictor of respiratory outcomes.

It is also worth noting that we are not considering the physiologic measures of apnea and periodic breathing (calculated by BAP using chest impedance waveform) in this analysis. These features are only meaningful for infants not on mechanical ventilation and have some of the same issues with respiratory support features mentioned above. They were also not shown to be major predictors of respiratory outcomes in primary Pre-Vent analysis (Ambalavanan et al. 2023).

## 3. Results

Figure 2 shows signature of risk curves for 5 of the top predictors of an unfavorable outcome at day 7. These are discussed in more detail below.

**Figure 2:**
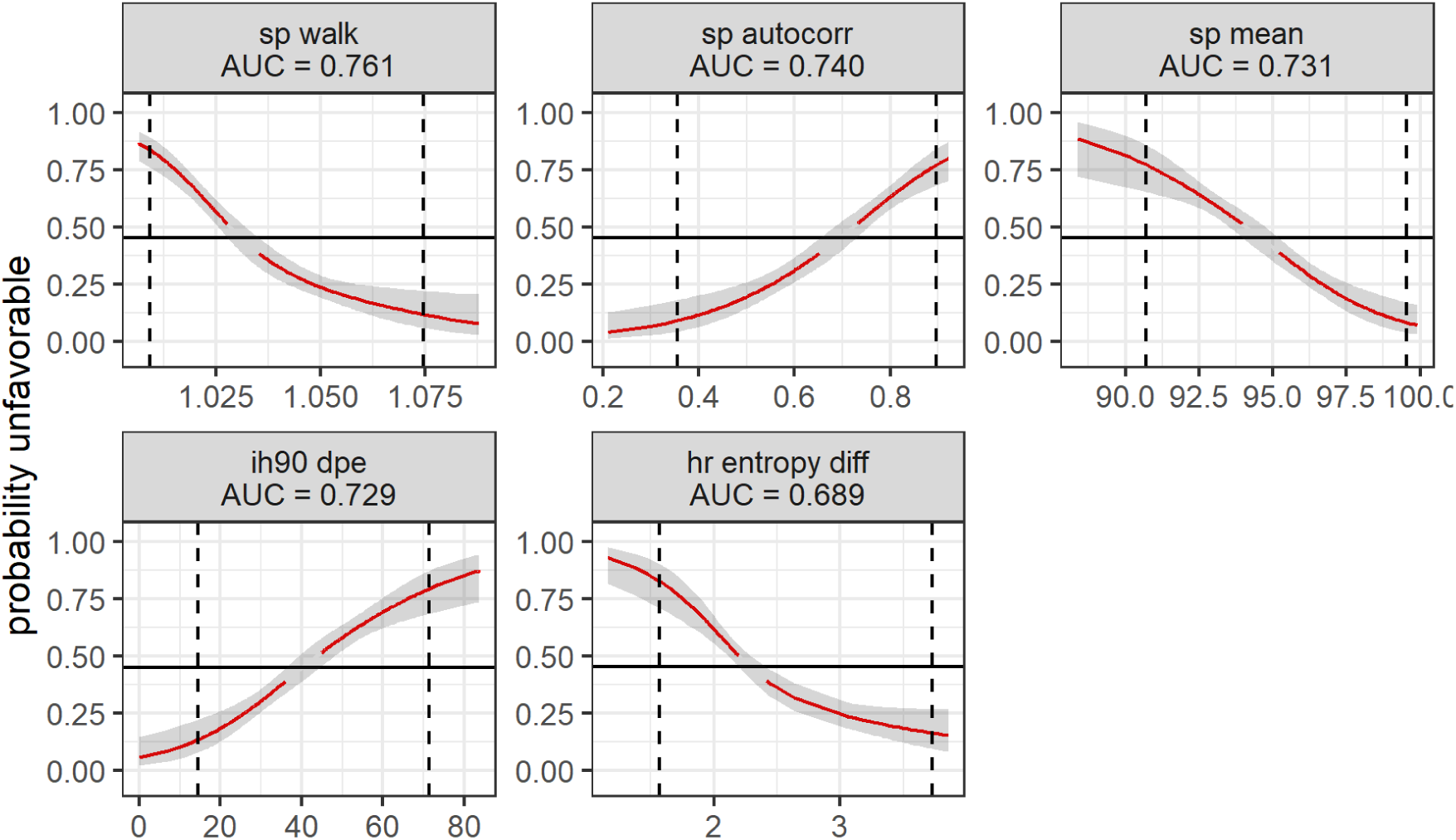
Individual signature of risk for unfavorable outcome for top predictors at Day 7. The gray area indicates the 95% confidence intervals. The black line represents the outcome rate of .454 and the red line indicates where the confidence intervals do not include this null value. The dotted lines represent the limits of the central 95% of the daily distribution values.

Table 3 summarizes the top individual performers in order of highest AUC for predicting primary unfavorable outcome at day 7. Results and ranks (out of 42) for days 14 and 28 are also included. Comprehensive results for all features and each of the outcomes for days 7, 14 and 28 is provided in Tables A2,A3, and A4. Performance of the major clinical risk factors are also provided in Table A5 for comparison.

**Table 3:**
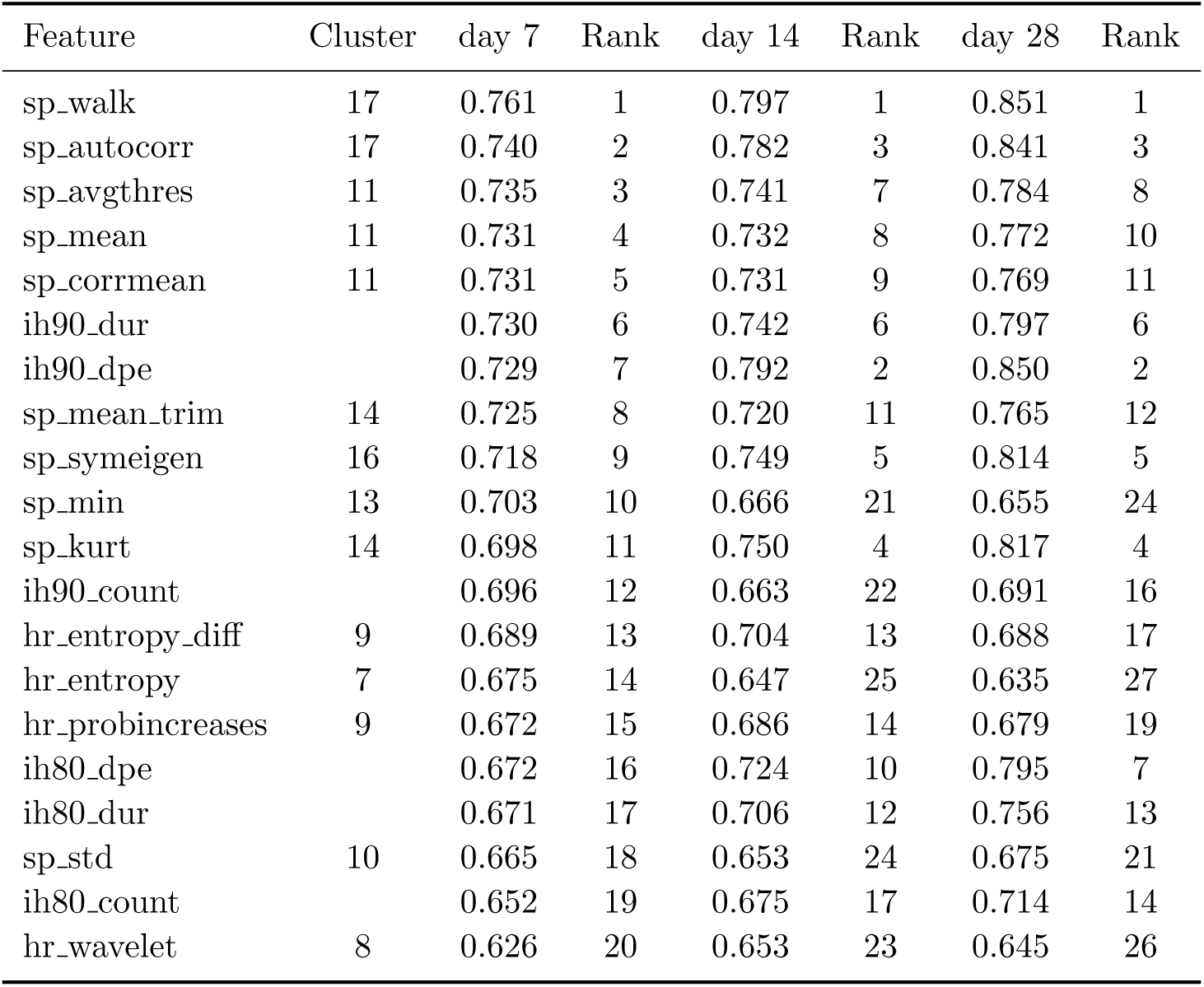
Top performers for predicting primary unfavorable respiratory outcome and respective ranks (out of 42) at day 7, 14 and 28.

### 3.1. Top SPO2 HCTSA Features

The top HCTSA feature for predicting respiratory outcomes was sp walk which is a time series metric inspired by physics. This may not be naturally intuitive for clinicians (and others) and perhaps not that useful despite its optimal performance. The feature is based on comparing the signal’s standard deviation to that of a simulated hypothetical particle (or ’walker’) that moves in response to values of the time series at each point. As described in the Matlab code, the walker moves as if it had inertia (based on its mass which is an input parameter) from the previous time step so that it ’wants’ to move the same amount and the original time series acts as a force changing its motion. SPO2 time series that behave very much like this simulated walk have lower ratios close to 1 which indicate higher risk of unfavorable outcomes.

This algorithm is part of a cluster of algorithms associated with the autocorrelation of the SPO2 time series and includes the central algorithm and second best performer sp_autocorr. Autocorrelation is a standard time domain tool to evaluate, among other things, the frequency content of the signal and as such likely a more intuitive approach than that of sp walk. The specific parameters for sp_autocorr was the correlation of SPO2 with a delayed copy of itself at a lag of 4 samples or 8 seconds. High values of Sp_autocorr are associated with unfavorable outcomes and occur in signals that have some variability over a 10-minute window but do not have high frequency variations locally (over an 8 second window). Looking more closely at other lags and metrics associated with the autocorrelation of the SPO2 is a potentially promising area for future work. Both sp_walk and sp_autocorr were highly correlated with IH90 DPE (*−.*77 and .76 respectively) and are fundamentally measuring the same physiological status of the infant but perhaps in a more robust way.

Figure 3 shows examples of 10-minute SPO2 time series from extremely premature infants on day of life 7 from publicly available dataset (Niestroy et al. 2021). Figure 3A and 3B shows the values and percentiles for sp walk and sp_autocorr respectively. The top row of each figure shows higher risk examples which is low values for sp walk and high values for sp_autocorr. These records are generally associated with persistent low-frequency patterns with desaturations that include long duration IH90 and IH80 events. The middle and bottom rows of these examples show median and percentiles associated with low risk. These exhibit some interesting patterns but not in a way that has consistent clinical interpretation like with high risk examples. Future work beyond the scope of this paper is needed to understand if the presence of subtle high-frequency variability in SPO2 is clinically meaningful.

**Figure 3:**
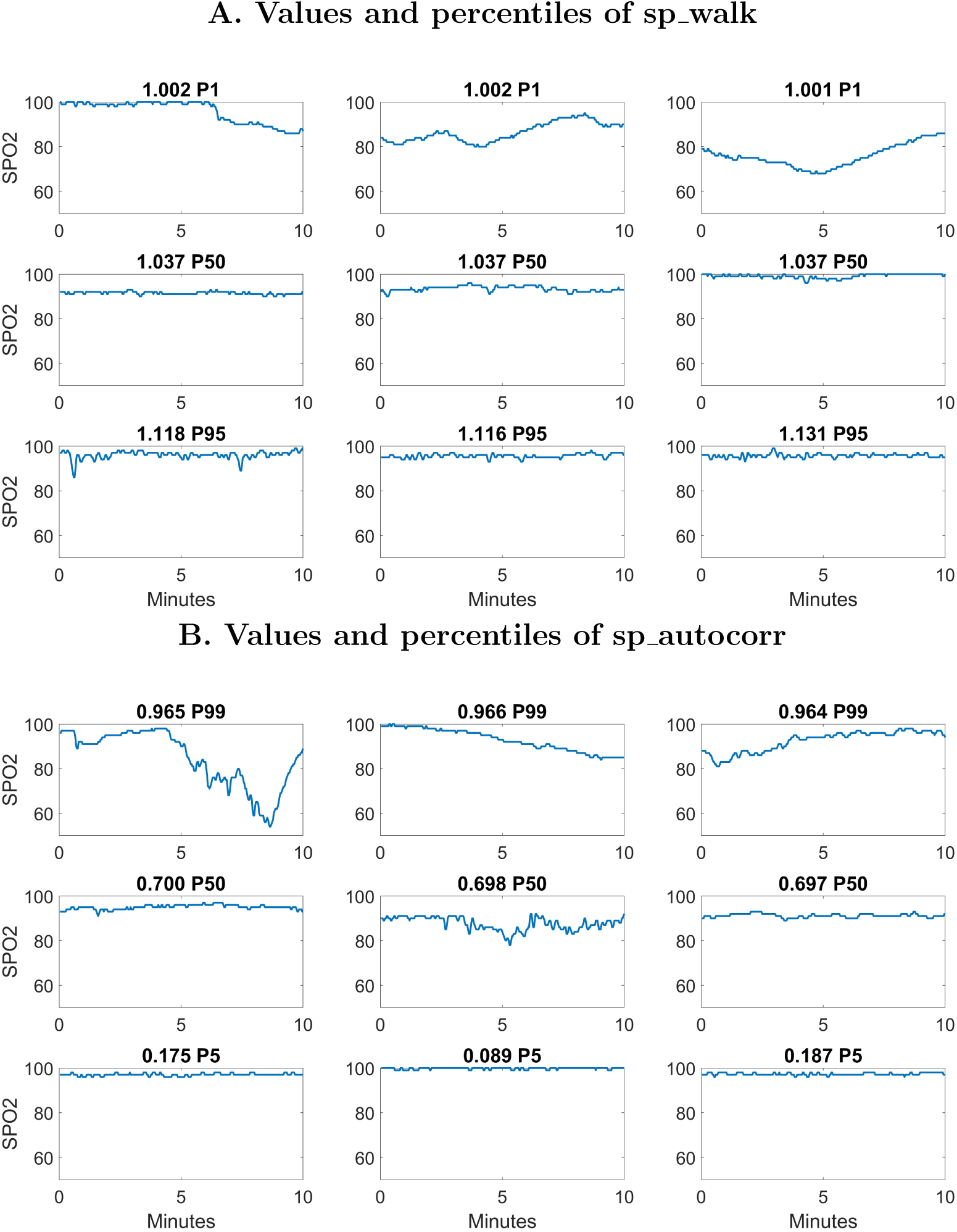
Examples of 10-minute SPO2 time series on day 7 for twp top HCTSA features sp walk and sp_autocorr. The title includes feature value and its percentile, e.g. P5=5*^th^* percentile of distribution. individual predictor of mortality in next week in the NICU (Niestroy et al. 2022). This measure transforms the HR record into binary sequence of either up (u) or down (d) based on whether HR increases or not (HR staying same is considered down). The hr probincreases is the probability of the uu pattern and low values indicate high risk of mortality.

Other top SPO2 features include sp mean and other measures from this cluster. None of these other features outperforms sp mean by an amount that would likely justify deviating from using a simple measure that is clinically easy to understand. Another SPO2 metric that performed well at days 14 and 28 was kurtosis. The mean and kurtosis are part of a larger group of SPO2 features that characterize a daily pulse oxygen histogram or profile. These histograms are used as a clinical tool in the NICU to evaluate target range management(Gentle et al. 2020, Goetz et al. 2022, Borenstein-Levin et al. 2020). Developing predictive models based directly on these daily SPO2 histograms is a promising area for future work. From an HCTSA perspective, this would correspond to restricting analysis to metrics that don’t depend on the order of the time series values.

### 3.2. Top HR HCTSA Features

The top three HR features were symbolic transformation metrics associated with discretizing the HR record into sequence of either 2 (binary) or 3 symbols that are also good measures of low HR variability associated with high risk of death. The best algorithm, hr entropy diff, specifically first quantizes the successive HR differences into 3 symbols roughly representing increasing, same and decreasing. It then looks at the distribution of all 3^4^ = 81 possible patterns of length 4 and calculates the Shannon entropy. Low values of hr entropy diff are associated with large proportion of unchanging HR during 10-minute window. The second best algorithm, hr entropy, is the exact same algorithm applied to raw signal without first taking differences. The third best HR algorithm is hr probincreases which was previously identified as best individual predictor of mortality in next week in the NICU (Niestroy et al. 2022). This measure transforms the HR record into binary sequence of either up (u) or down (d) based on whether HR increases or not (HR staying same is considered down). The hr probincreases is the probability of the uu pattern and low values indicate high risk of mortality.

All three of these measures are related to heart rate fragmentation metrics introduced recently (Costa et al. 2017*a*,*b*) and shown to be predictive of long-term survival in large study of 3000 24 hour Holter recordings from patients of all ages (Lensen et al. 2020). A lesson learned from HCTSA is that measures that convert the HR to a simple binary time series and look for runs of ones of lengths 1,2,3,. . . can quantify heart-rate variability and are good candidates to predict adverse neonatal outcomes in a way clinicians can easily understand.

### 3.3. Trajectories of Predictive Performance

Values of the AUC for sp walk and IH90 DPE at days 7, 14 and 28 are summarized in Table 4. Based on the individual results discussed above, a simple 3-parameter HCTSA model using features sp walk, sp mean, and hr entropy diff was developed at each day of life and also included. This model was considered sufficiently optimal because removing any of these features reduced AUC by more than .005 and no HCTSA feaure added to the model increased AUC by more than .005 at day 7. The performance of birth weight alone is included as well for comparison.

**Table 4:** Comparison of AUC at days 7, 14 and 28 for predicting respiratory outcomes. The HCTSA model used the 3 features sp walk, sp mean, and hr entropy diff.

| Features | Day | unfavorable | moderate/severe/death | severe/death | death |
| --- | --- | --- | --- | --- | --- |
| ih90_dpe | 7 | 0.729 | 0.715 | 0.731 | 0.719 |
| sp_walk | 7 | 0.761 | 0.724 | 0.734 | 0.675 |
| hctsa model | 7 | 0.778 | 0.754 | 0.757 | 0.747 |
| birth weight | 7 | 0.798 | 0.788 | 0.786 | 0.801 |
| ih90_dpe | 14 | 0.791 | 0.767 | 0.761 | 0.741 |
| sp_walk | 14 | 0.797 | 0.763 | 0.738 | 0.689 |
| hctsa model | 14 | 0.800 | 0.777 | 0.762 | 0.765 |
| birth weight | 14 | 0.798 | 0.787 | 0.781 | 0.808 |
| ih90_dpe | 28 | 0.850 | 0.866 | 0.843 | 0.770 |
| sp_walk | 28 | 0.851 | 0.860 | 0.834 | 0.779 |
| hctsa model | 28 | 0.849 | 0.857 | 0.831 | 0.758 |
| birth weight | 28 | 0.792 | 0.801 | 0.802 | 0.849 |

The AUC of the models in Table 4 were calculated for each respiratory outcome from birth up to 8 weeks of life. Figure 4 shows these performance trajectory curves.

**Figure 4:**
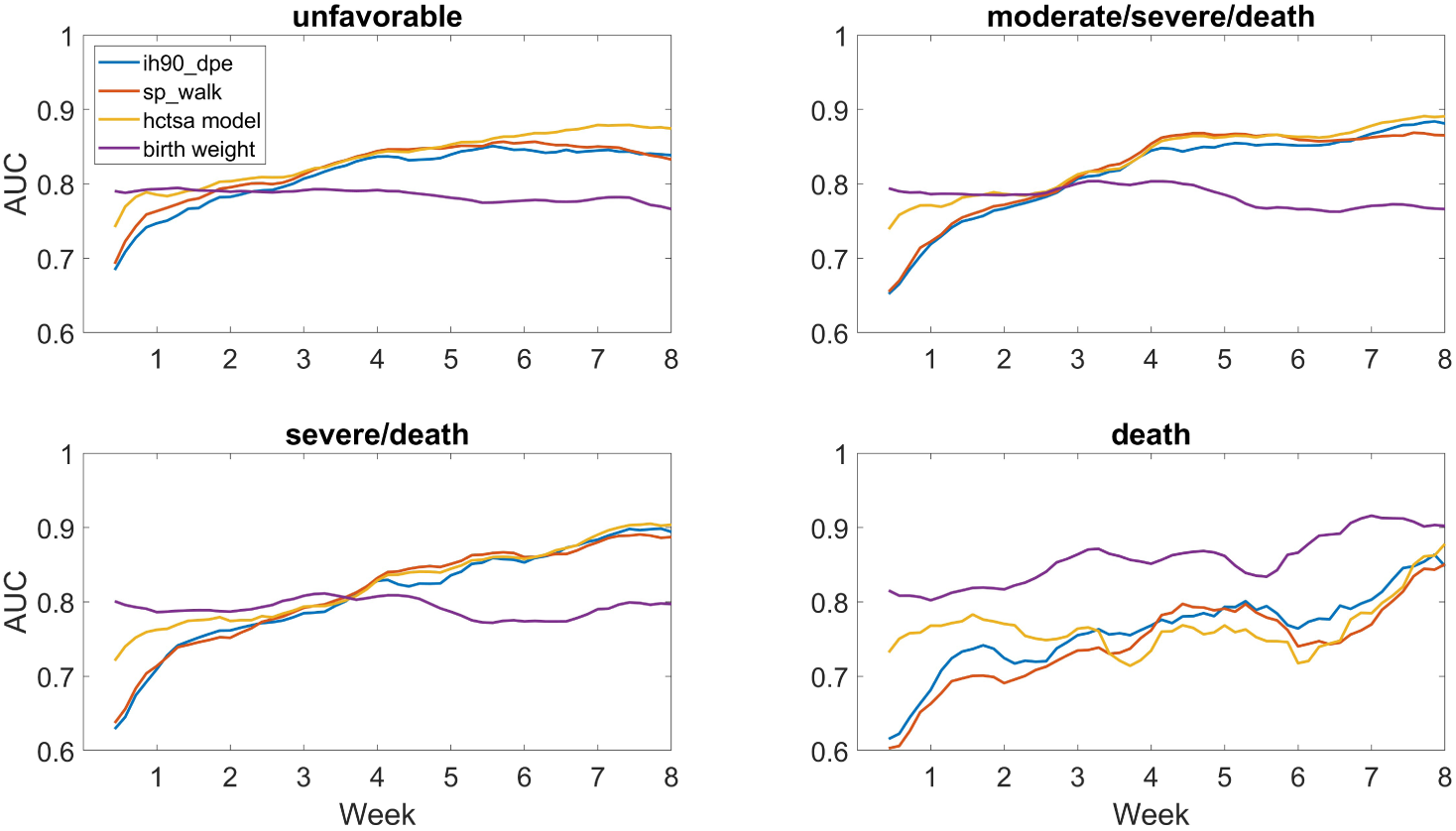
Trajectory of AUC by week of life for predicting respiratory outcomes. Results are averaged over *±* 2 days.

The evolving variable importance of each of the 3 features in the HCTSA model is shown in Figure 5. All these curves are smoothed by taking average over window of plus or minus two days. Detailed trajectories of the physiologic features including IH events are available for the Pre-Vent cohort (Weese-Mayer et al. 2023).

**Figure 5:**
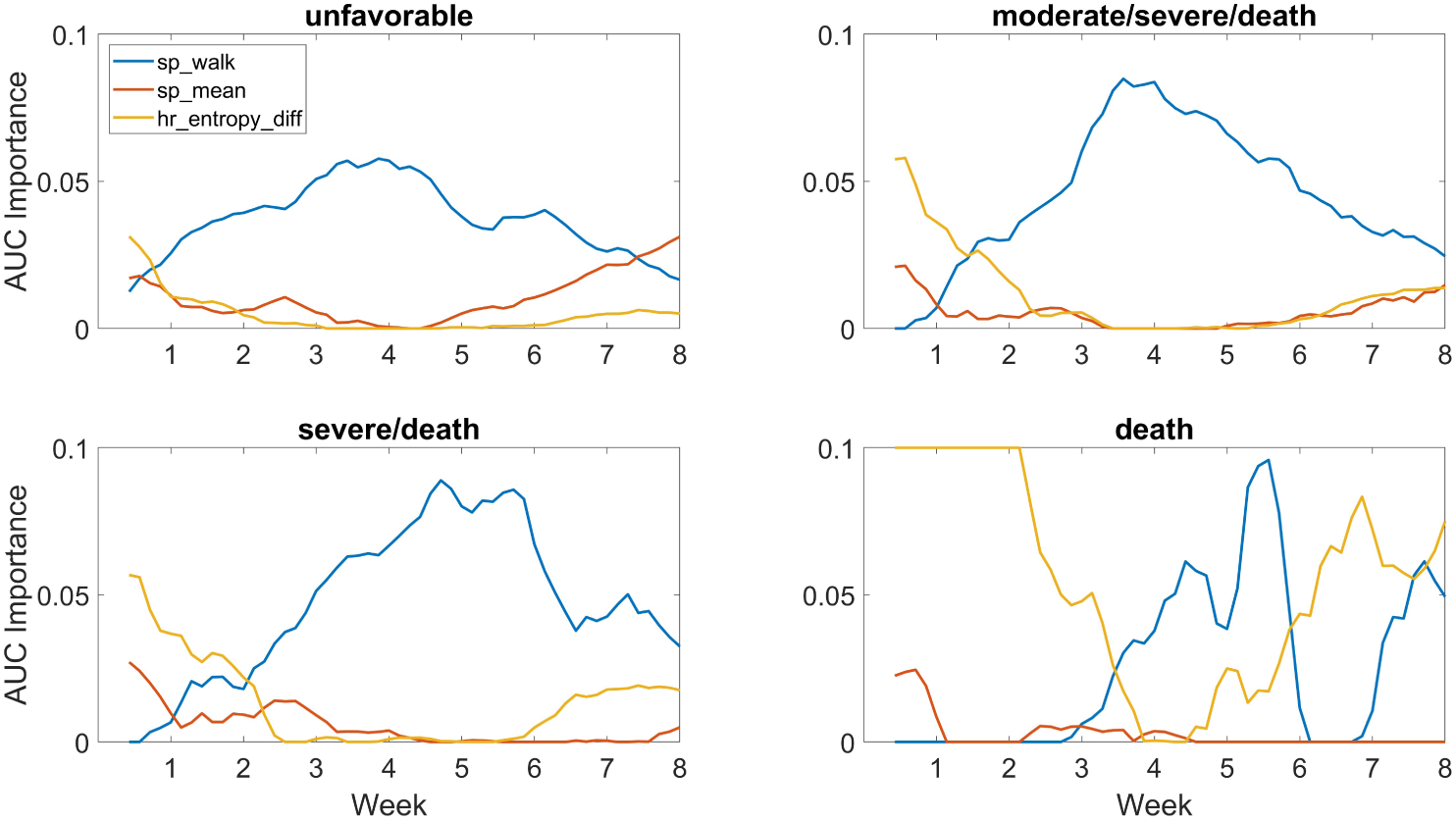
Importance of HCTSA features by week of life measured by drop in AUC when variable is dropped from 3-parameter model. Results are averaged over *±* 2 days.

For unfavorable outcome, the top HCTSA feature slightly outperformed IH90 DPE at day 7 (.761 to .729) but was essentially equivalent at day 28 (.851 to .850). The HCTSA model increased the AUC to .778 at day 7 but did not improve performance at day 28 (.849). Models with new HCTSA features still have a lower AUC than birth weight alone at day 7, but catch up and surpass birth weight by day 14. By week 4 or so, all the physiologic models outperform birth weight for predicting all outcomes except death.

The variable importance of the IH related HCTSA feature sp walk steadily increases to day of life 28 where it is essentially the only predictor needed. The top HR metric was only an important predictor at early ages which was likely due to the larger proportion of infants whose bad outcome is death by any cause.

## 4. Discussion

A representative subset of HCTSA algorithms predicted unfavorable respiratory outcomes in extremely preterm infants and revealed both new robust features associated with IH events and other features for improved detection early in NICU stay. This large multi-center cohort study validated the hypothesis that identifying algorithms by unsupervised clustering could capture most of the predictive information in NICU HR and SPO2 data. However, this analysis also showed that HCTSA did not outperform IH metrics later in the NICU stay and provides additional evidence supporting IH90 DPE as an optimal predictor of respiratory outcomes.

As with all metrics based on bedside monitoring data, there are significant practical issues associated with implementation especially for wide-spread clinical use. The data used in the NICU study to perform the unsupervised clustering used in this paper was from HR and SPO2 displayed on bedside monitors every 2 seconds. The PreVent sites had a variety of bedside monitors that displayed vital signs either once every second or every 1024 milliseconds. To accommodate for this, data was sub-sampled every 2 seconds at all sites before applying algorithms. For the analysis of HR, one way to minimize variability across sites is to work directly with the interbeat RR intervals derived from ECG waveforms. However, this often involves a significant amount of additional data processing and management that may not be practical. As mentioned previously, SPO2 metrics including IH events can also be sensitive to the averaging time of the pulse oximeter which can vary across vendor and NICU.

In this study, the predictive performance of the features were analyzed at a daily level to be consistent with the PreVent study database and primary analysis. Future work would be to look at higher resolution windows of hourly or even down to individual 10-minute records. This would also involve a closer look at individual IH events and their correlation with sp walk and sp_autocorr.

One possible limitation of this study was that the algorithms implemented in (Niestroy et al. 2022) did not include the entire HCTSA algorithm library of over 7000 features because it was not computationally feasible to implement on such a large data set. It is believed that the algorithms implemented were sufficient to be a reasonable implementation of HCTSA approach and extremely useful in developing predictive models in NICU. Also it was not feasible to implement a much larger set of HCTSA algorithms on the Pre-Vent cohort since the data was spread out among 5 sites which led to only calculating the 33 features. Augmenting the BAP to efficiently compute a larger number of physiologic features to help address these issues will be included in future analyses of the Pre-Vent cohort.

## Data Availability

The datasets are available from the corresponding author upon reasonable request.

## 5. Acknowledgements

The National Institutes of Health and the National Heart, Lung, and Blood Institute (NHLBI) provided grant support through cooperative agreements. While NHLBI staff did have input into the study design, conduct, analysis, and manuscript drafting, the content and views expressed are solely the responsibility of the authors and do not necessarily represent the official views of the National Institutes of Health or the U.S. Department of Health and Human Services.

Participating sites collected and stored the data while the University of Virginia, the lead data coordinating center (LDCC), analyzed the data. The co-PIs at each site had full access to his/her individual site data and take responsibility for the integrity of the raw waveforms while Drs. Randall Moorman (LDCC co-PI) and Douglas Lake (LDCC co-PI) take responsibility for the integrity of the data and accuracy of the data analysis.

We are indebted to our medical and nursing colleagues and the infants and their parents who agreed to take part in this study. The following investigators, in addition to those listed as authors, participated in this study:

## Pre-Vent Investigators

Katy N. Krahn and Amanda M. Zimmet, University of Virginia Center for Advanced Medical Analytics, Charlottesville, VA; Bradley S Hopkins, Erin K Lonergan, Casey M. Rand: Ann and Robert H. Lurie Children’s Hospital of Chicago, and Pediatric Autonomic Medicine, Stanley Manne Children’s Research Institute, Chicago, IL; Arlene Zadell, Department of Pediatrics, University Hospitals Cleveland Medical Center, Rainbow Babies and Children’s Hospital, Cleveland, OH; Arie Nakhmani, Department of Electrical and Computer Engineering, University of Alabama at Birmingham, Birmingham, AL; Waldemar A. Carlo, Deborah Laney, and Colm P. Travers, Division of Neonatology, Department of Pediatrics, University of Alabama at Birmingham School of Medicine, Birmingham, AL; Silvia Vanbuskirk, Carmen D’Ugard, Ana Cecilia Aguilar and Alini Schott, Division of Neonatology, Department of Pediatrics, University of Miami Miller School of Medicine, Holtz Children’s Hospital – University of Miami/Jackson Memorial Medical Center, Miami, FL; and Julie Hoffmann and Laura Linneman, Washington University School of Medicine in St. Louis, St. Louis, MO.

## NIH/NHLBI

Neil Aggarwal, MD: National Institutes of Health, National Heart, Lung and Blood Institute, Division of Lung Diseases, Bethesda, MD

Lawrence Baizer, PhD: National Institutes of Health, National Heart, Lung and Blood Institute, Division of Lung Diseases, Bethesda, MD

Peyvand Ghofrani, MDE: National Institutes of Health, National Heart, Lung and

Blood Institute, Division of Lung Diseases, Bethesda, MD Aaron D. Laposky, PhD: National Institutes of Health, National Heart, Lung and Blood Institute, Division of Lung Diseases, Bethesda, MD

Aruna Natarajan MD, PhD: National Institutes of Health, National Heart, Lung and Blood Institute, Division of Lung Diseases, Bethesda, MD

Barry Schmetter, BS: National Institutes of Health, National Heart, Lung and Blood Institute, Division of Lung Diseases, Bethesda, MD

## OSMB

Estelle B. Gauda, MD (Chair): University of Toronto Hospital for Sick Children, Division of Neonatology, Toronto, Ontario

Jonathan M. Davis, MD: Tufts Clinical and Translational Science Institute, Division of Newborn Medicine, Boston, MA

Roberta L. Keller, MD: University of California, San Francisco School of Medicine, Department of Pediatrics, San Francisco CA

Robinder G. Khemani, MD: Children’s Hospital Los Angeles, Department of Anesthesiology and Critical Care Medicine, Los Angeles, CA

Renee H. Moore, PhD: Drexel University Department of Epidemiology and Biostatistics, Philadelphia, PA

Elliott M. Weiss, MD, MSME: Seattle Children’s Hospital, Department of Pediatrics, Division of Bioethics, Seattle, WA

## Funding Support

University of Virginia (NCT03174301): U01 HL133708, K23 HD097254, HL133708-05S1

Case Western Reserve University: U01 HL133643 Northwestern University: U01 HL133704

University of Alabama at Birmingham: U01 HL133536, K23 HL157618 University of Miami: U01 HL133689

Washington University: U01 HL133700, K23 NS111086

We give a special acknowledgement to Amanda Zimmet whose work developing the Batch Algorithm Processor was crucial to this work and success of the Pre-Vent study. We also thank Alix Zimmet for his acknowledgement skills and raising the bar for manuscript preparation.

## Appendix A. Supplemental tables

**Table A1:**
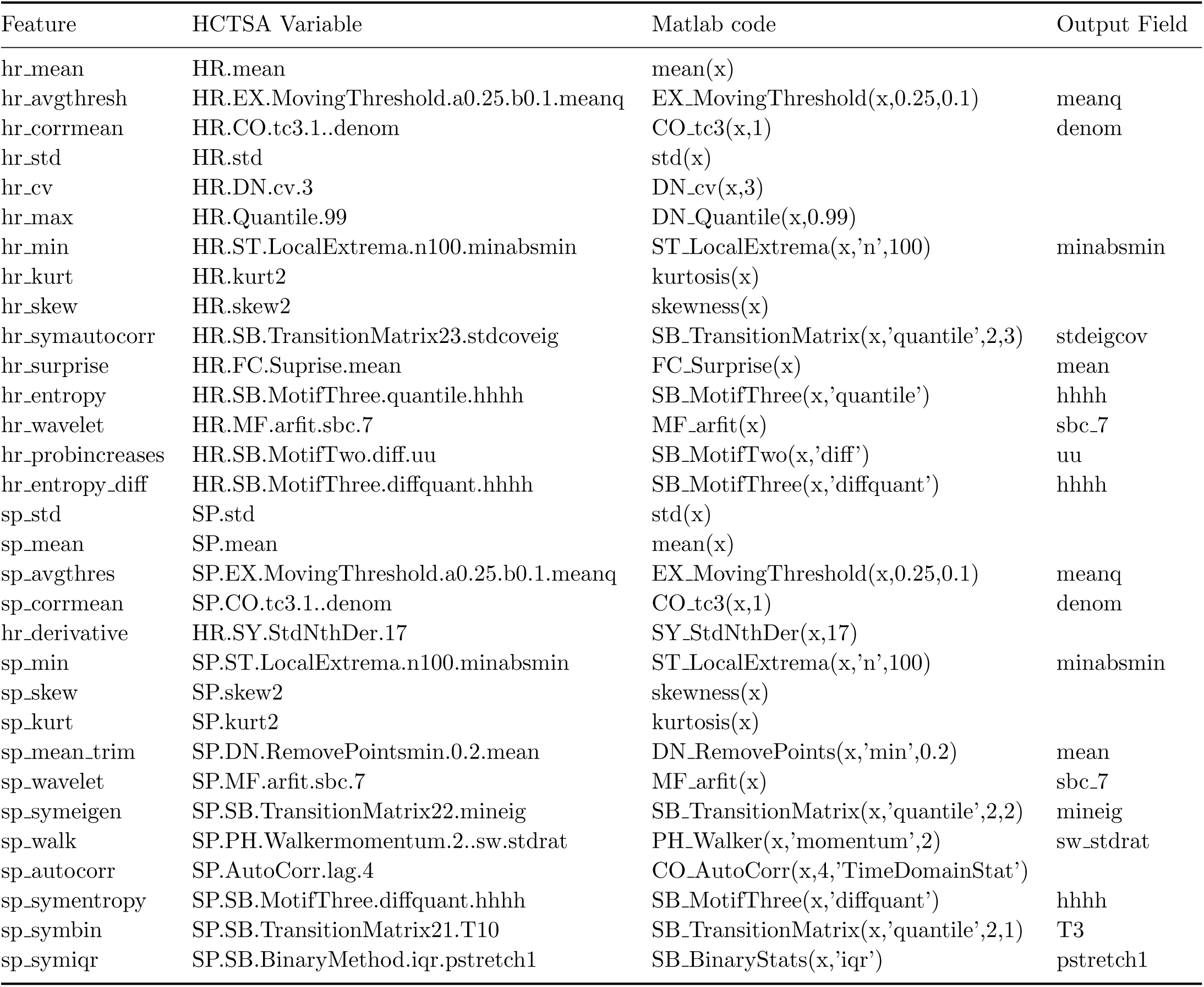
HCTSA variable names and Matlab functions available to download from https://github.com/benfulcher/hctsa.

| Feature | HCTSA Variable | Matlab code | Output Field |
| --- | --- | --- | --- |
| hr_mean | HR.mean | mean(x) |  |
| hr_avgthresh | HR.EX.MovingThreshold.a0.25.b0.1.meanq | EX_MovingThreshold(x,0.25,0.1) | meanq |
| hr_corrmean | HR.CO.tc3.1..denom | CO_tc3(x,1) | denom |
| hr_std | HR.std | std(x) |  |
| hr_cv | HR.DN.cv.3 | DN_cv(x,3) |  |
| hr_max | HR.Quantile.99 | DN_Quantile(x,0.99) |  |
| hr_min | HR.ST.LocalExtrema.n100.minabsmin | ST_LocalExtrema(x,'n',100) | minabsmin |
| hr_kurt | HR.kurt2 | kurtosis(x) |  |
| hr_skew | HR.skew2 | skewness(x) |  |
| hr_symautocorr | HR.SB.TransitionMatrix23.stdcoveig | SB_TransitionMatrix(x,'quantile',2,3) | stdeigcov |
| hr_surprise | HR.FC.Surprise.mean | FC_Surprise(x) | mean |
| hr_entropy | HR.SB.MotifThree.quantile.hhhh | SB_MotifThree(x,'quantile') | hhhh |
| hr_wavelet | HR.MF.arfit.sbc.7 | MF_arfit(x) | sbc_7 |
| hr_probincreases | HR.SB.MotifTwo.diff.uu | SB_MotifTwo(x,'diff') | uu |
| hr_entropy_diff | HR.SB.MotifThree.diffquant.hhhh | SB_MotifThree(x,'diffquant') | hhhh |
| sp_std | SP.std | std(x) |  |
| sp_mean | SP.mean | mean(x) |  |
| sp_avgthres | SP.EX.MovingThreshold.a0.25.b0.1.meanq | EX_MovingThreshold(x,0.25,0.1) | meanq |
| sp_corrmean | SP.CO.tc3.1..denom | CO_tc3(x,1) | denom |
| hr_derivative | HR.SY.StdNthDer.17 | SY_StdNthDer(x,17) |  |
| sp_min | SP.ST.LocalExtrema.n100.minabsmin | ST_LocalExtrema(x,'n',100) | minabsmin |
| sp_skew | SP.skew2 | skewness(x) |  |
| sp_kurt | SP.kurt2 | kurtosis(x) |  |
| sp_mean_trim | SP.DN.RemovePointsmin.0.2.mean | DN_RemovePoints(x,'min',0.2) | mean |
| sp_wavelet | SP.MF.arfit.sbc.7 | MF_arfit(x) | sbc_7 |
| sp_symeigen | SP.SB.TransitionMatrix22.mineig | SB_TransitionMatrix(x,'quantile',2,2) | mineig |
| sp_walk | SP.PH.Walkermomentum.2..sw.stdrat | PH_Walker(x,'momentum',2) | sw_stdrat |
| sp_autocorr | SP.AutoCorr.lag.4 | CO_AutoCorr(x,4,'TimeDomainStat') |  |
| sp_symentropy | SP.SB.MotifThree.diffquant.hhhh | SB_MotifThree(x,'diffquant') | hhhh |
| sp_symbin | SP.SB.TransitionMatrix21.T10 | SB_TransitionMatrix(x,'quantile',2,1) | T3 |
| sp_symiqr | SP.SB.BinaryMethod.iqr.pstretch1 | SB_BinaryStats(x,'iqr') | pstretch1 |

**Table A2:** AUC at day 7 of all individual HR and SPO2 features on increasingly bad respiratory outcomes.

| Feature | Cluster | unfavorable | moderate/severe/death | severe/death | death |
| --- | --- | --- | --- | --- | --- |
| hr_mean | 1 | 0.546 | 0.564 | 0.576 | 0.624 |
| hr_corrmean | 1 | 0.547 | 0.566 | 0.578 | 0.626 |
| hr_avgthresh | 1 | 0.554 | 0.573 | 0.587 | 0.638 |
| hr_std | 2 | 0.623 | 0.643 | 0.666 | 0.693 |
| hr_cv | 2 | 0.613 | 0.629 | 0.651 | 0.668 |
| hr_max | 3 | 0.588 | 0.610 | 0.628 | 0.674 |
| hr_min | 4 | 0.490 | 0.500 | 0.493 | 0.517 |
| hr_skew | 5 | 0.550 | 0.540 | 0.526 | 0.492 |
| hr_kurt | 5 | 0.506 | 0.494 | 0.532 | 0.534 |
| hr_symautocorr | 6 | 0.610 | 0.578 | 0.555 | 0.630 |
| hr_surprise | 7 | 0.624 | 0.625 | 0.672 | 0.718 |
| hr_entropy | 7 | 0.675 | 0.673 | 0.665 | 0.736 |
| hr_wavelet | 8 | 0.626 | 0.635 | 0.649 | 0.689 |
| hr_entropy_diff | 9 | 0.689 | 0.706 | 0.709 | 0.765 |
| hr_probincreases | 9 | 0.672 | 0.688 | 0.702 | 0.744 |
| sp_std | 10 | 0.665 | 0.641 | 0.598 | 0.560 |
| sp_mean | 11 | 0.731 | 0.694 | 0.692 | 0.670 |
| sp_corrmean | 11 | 0.731 | 0.693 | 0.692 | 0.669 |
| sp_avgthresh | 11 | 0.735 | 0.699 | 0.700 | 0.678 |
| hr_derivative | 12 | 0.601 | 0.598 | 0.611 | 0.651 |
| sp_min | 13 | 0.703 | 0.687 | 0.671 | 0.635 |
| sp_skew | 14 | 0.614 | 0.587 | 0.559 | 0.601 |
| sp_kurt | 14 | 0.698 | 0.646 | 0.614 | 0.608 |
| sp_mean_trim | 14 | 0.725 | 0.686 | 0.688 | 0.667 |
| sp_wavelet | 15 | 0.540 | 0.503 | 0.549 | 0.570 |
| sp_symeigen | 16 | 0.718 | 0.683 | 0.648 | 0.609 |
| sp_autocorr | 17 | 0.740 | 0.702 | 0.692 | 0.645 |
| sp_walk | 17 | 0.761 | 0.724 | 0.734 | 0.675 |
| sp_symentropy | 18 | 0.552 | 0.509 | 0.538 | 0.548 |
| sp_symbin | 19 | 0.595 | 0.614 | 0.626 | 0.618 |
| sp_symiqr | 20 | 0.600 | 0.626 | 0.636 | 0.626 |
| xc_max |  | 0.511 | 0.559 | 0.577 | 0.622 |
| xc_min |  | 0.538 | 0.571 | 0.595 | 0.636 |
| ih90_count |  | 0.696 | 0.679 | 0.637 | 0.604 |
| ih90_dur |  | 0.730 | 0.715 | 0.691 | 0.658 |
| ih90_dpe |  | 0.729 | 0.715 | 0.731 | 0.719 |
| ih80_count |  | 0.652 | 0.673 | 0.666 | 0.678 |
| ih80_dur |  | 0.671 | 0.692 | 0.688 | 0.704 |
| ih80_dpe |  | 0.672 | 0.685 | 0.677 | 0.710 |
| brady_count |  | 0.612 | 0.631 | 0.613 | 0.542 |
| brady_dur |  | 0.595 | 0.615 | 0.595 | 0.517 |
| brady_dpe |  | 0.542 | 0.584 | 0.564 | 0.508 |

**Table A3:**
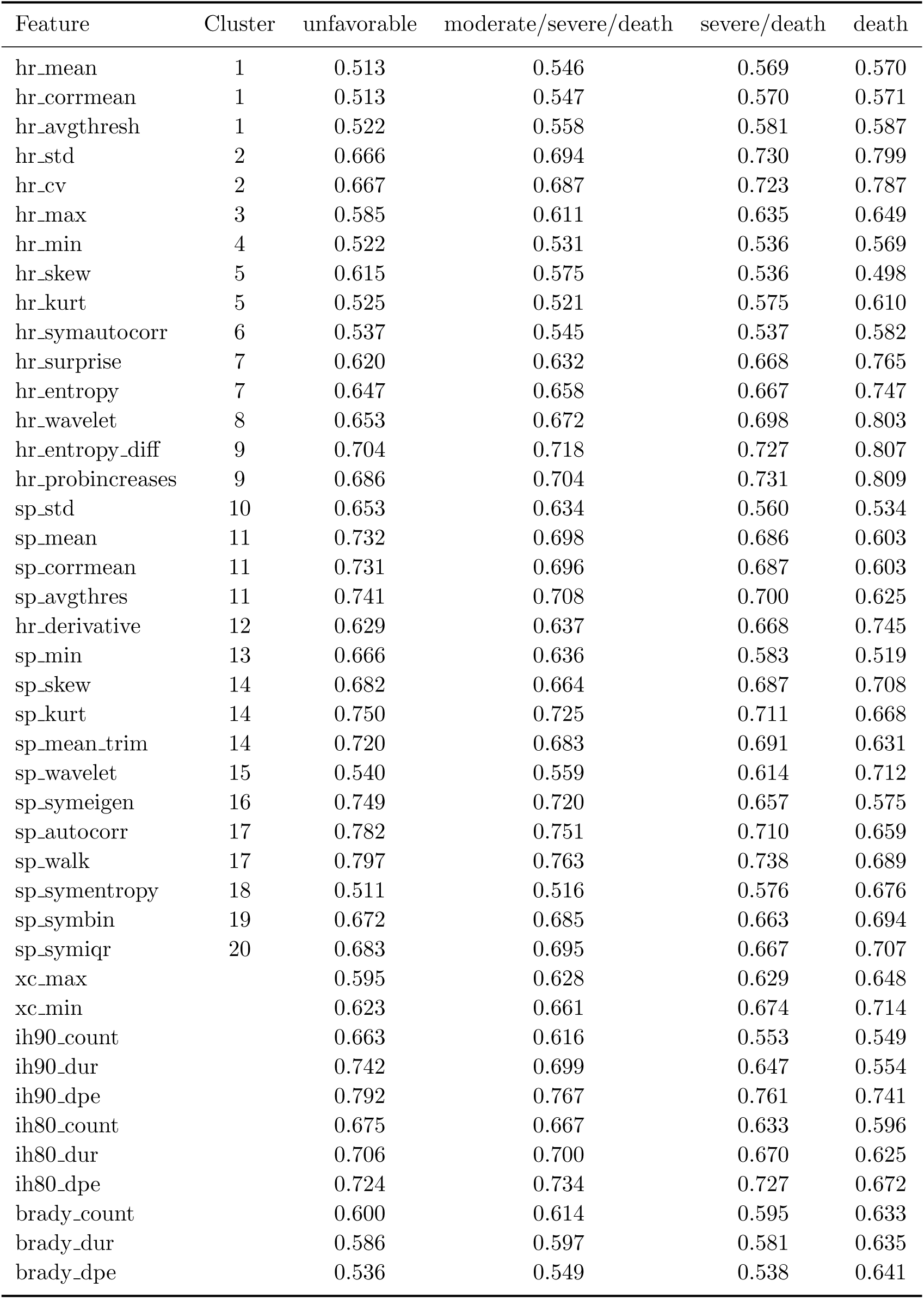
AUC at day 14 of all individual HR and SPO2 features on increasingly bad respiratory outcomes.

**Table A4:** AUC at day 28 of all individual HR and SPO2 features on increasingly bad respiratory outcomes.

| Feature | Cluster | unfavorable | moderate/severe/death | severe/death | death |
| --- | --- | --- | --- | --- | --- |
| hr_mean | 1 | 0.524 | 0.588 | 0.608 | 0.581 |
| hr_corrmean | 1 | 0.525 | 0.589 | 0.608 | 0.580 |
| hr_avgthresh | 1 | 0.533 | 0.600 | 0.620 | 0.589 |
| hr_std | 2 | 0.657 | 0.679 | 0.696 | 0.678 |
| hr_cv | 2 | 0.651 | 0.655 | 0.669 | 0.656 |
| hr_max | 3 | 0.604 | 0.664 | 0.681 | 0.657 |
| hr_min | 4 | 0.515 | 0.549 | 0.559 | 0.569 |
| hr_skew | 5 | 0.679 | 0.661 | 0.641 | 0.584 |
| hr_kurt | 5 | 0.558 | 0.556 | 0.541 | 0.505 |
| hr_symautocorr | 6 | 0.544 | 0.572 | 0.552 | 0.552 |
| hr_surprise | 7 | 0.663 | 0.673 | 0.708 | 0.802 |
| hr_entropy | 7 | 0.635 | 0.642 | 0.626 | 0.673 |
| hr_wavelet | 8 | 0.645 | 0.679 | 0.694 | 0.685 |
| hr_entropy_diff | 9 | 0.688 | 0.697 | 0.705 | 0.719 |
| hr_probincreases | 9 | 0.679 | 0.702 | 0.715 | 0.751 |
| sp_std | 10 | 0.675 | 0.620 | 0.588 | 0.492 |
| sp_mean | 11 | 0.772 | 0.754 | 0.748 | 0.768 |
| sp_corrmean | 11 | 0.769 | 0.753 | 0.748 | 0.770 |
| sp_avgthres | 11 | 0.784 | 0.771 | 0.766 | 0.790 |
| hr_derivative | 12 | 0.612 | 0.625 | 0.642 | 0.378 |
| sp_min | 13 | 0.655 | 0.595 | 0.577 | 0.543 |
| sp_skew | 14 | 0.783 | 0.779 | 0.767 | 0.752 |
| sp_kurt | 14 | 0.817 | 0.821 | 0.799 | 0.745 |
| sp_mean_trim | 14 | 0.765 | 0.750 | 0.759 | 0.790 |
| sp_wavelet | 15 | 0.577 | 0.652 | 0.647 | 0.667 |
| sp_symeigen | 16 | 0.814 | 0.794 | 0.750 | 0.691 |
| sp_autocorr | 17 | 0.841 | 0.844 | 0.807 | 0.728 |
| sp_walk | 17 | 0.851 | 0.860 | 0.834 | 0.779 |
| sp_symentropy | 18 | 0.532 | 0.543 | 0.554 | 0.621 |
| sp_symbin | 19 | 0.681 | 0.706 | 0.687 | 0.673 |
| sp_symiqr | 20 | 0.704 | 0.728 | 0.705 | 0.687 |
| xc_max |  | 0.606 | 0.652 | 0.648 | 0.590 |
| xc_min |  | 0.634 | 0.673 | 0.665 | 0.616 |
| ih90_count |  | 0.691 | 0.618 | 0.595 | 0.580 |
| ih90_dur |  | 0.797 | 0.760 | 0.737 | 0.680 |
| ih90_dpe |  | 0.850 | 0.866 | 0.843 | 0.770 |
| ih80_count |  | 0.714 | 0.669 | 0.655 | 0.604 |
| ih80_dur |  | 0.756 | 0.727 | 0.727 | 0.646 |
| ih80_dpe |  | 0.795 | 0.798 | 0.816 | 0.711 |
| brady_count |  | 0.542 | 0.541 | 0.468 | 0.430 |
| brady_dur |  | 0.475 | 0.481 | 0.500 | 0.429 |
| brady_dpe |  | 0.502 | 0.510 | 0.525 | 0.590 |

**Table A5:** AUC at day 7, 14 and day 28 of individual clinical features on increasingly bad respiratory outcomes.

| Feature | Day | unfavorable | moderate/severe/death | severe/death | death |
| --- | --- | --- | --- | --- | --- |
| birth weight | 7 | 0.798 | 0.788 | 0.786 | 0.801 |
| gestational age | 7 | 0.765 | 0.794 | 0.765 | 0.782 |
| fio2 | 7 | 0.763 | 0.733 | 0.736 | 0.696 |
| imv | 7 | 0.736 | 0.755 | 0.745 | 0.720 |
| birth weight | 14 | 0.787 | 0.780 | 0.781 | 0.808 |
| gestational age | 14 | 0.770 | 0.779 | 0.748 | 0.780 |
| fio2 | 14 | 0.797 | 0.783 | 0.789 | 0.765 |
| imv | 14 | 0.748 | 0.770 | 0.752 | 0.734 |
| birth weight | 28 | 0.792 | 0.801 | 0.802 | 0.849 |
| gestational age | 28 | 0.776 | 0.807 | 0.776 | 0.833 |
| fio2 | 28 | 0.829 | 0.817 | 0.791 | 0.754 |
| imv | 28 | 0.737 | 0.789 | 0.777 | 0.722 |

